# Long-read DNA and RNA sequencing reveal an intronic retrotransposon insertion in *TCOF1* causing Treacher Collins syndrome

**DOI:** 10.1101/2025.04.24.25326319

**Authors:** Federico Ferraro, Nikolas Kühn, Dmitrijs Rots, Herma C. van der Linde, Banin Mohseni, Leontine van Unen, Mark Drost, Mark Nellist, Marieke Koekkoek, Rachel Schot, Henriette W. de Gier, Mieke Pleumeekers, Tahsin Stefan Barakat, Tjitske Kleefstra, Marjolein Weerts, Marieke F. van Dooren, Tjakko J. van Ham

## Abstract

Treacher Collins syndrome (TCS) is a craniofacial genetic disorder caused by loss of function variants in *TCOF1, POLR1B, POLR1C* or *POLR1D*. Here we describe two previously undiagnosed half-siblings affected with clinical TCS, and their apparently unaffected parent. Diagnostic short-read RNA-Sequencing identified aberrant expression of *TCOF1* and optical genome mapping detected a large genomic insertion therein. Long-read genome sequencing (LR-GS) resolved a deep intronic 3.5 kb SINE-VNTR-Alu (SVA) retrotransposon insertion in intron 17 of *TCOF1*. Long read RNA-Seq demonstrated that the insertion was partially exonized inducing isoform switch to the shorter non-canonical *TCOF1* isoform c. SVA-insertion was confirmed in both half-siblings and we detected mosaicism one paarent. This is the first description of a retrotransposon causing TCS, and the first intronic SVA causing isoform switch as a disease mechanism. This work demonstrates the potential of LR-RNA-Seq and LR-GS, to identify pathogenic variants in unexplained genetic disorders.

## Introduction

Treacher Collins syndrome (TCS) is a rare congenital genetic disorder affecting craniofacial development caused in 89% of the patients by pathogenic variant in *TCOF1* (OMIM: 606847)^1^. *TCOF1* encodes a nucleolar protein that plays a crucial role in the synthesis of ribosomal RNA (rRNA) and proper craniofacial development^1^. *TCOF1* haploinsufficiency leads to cell-autonomous defects in neural crest cell formation and proliferation ^2^. To date, over 200 autosomal dominant (AD) pathogenic variants in *TCOF1* have been reported ^1^, typically leading to the production of a truncated and/or non-functional protein. In addition to pathogenic AD variants in *TCOF1*, a minority of TCS individuals result from pathogenic variants in *POLR1B (*AD), or pathogenic autosomal recessive variants (AR) in *POLR1C*, or *POLR1D (*AD or AR). These genes encode subunits of RNA-polymerase I, which is required for rDNA transcription, converging on the same mechanism as *TCOF1*. It is estimated that in ~4% of typical TCS a molecular cause cannot be identified, suggesting either the presence of a pathogenic variant in one of the known genes undetected by current technology, or possibly in another gene not currently linked to TCS ^3^.

Here, we report an unconventional molecular genetic mechanism causing *TCOF1* haploinsufficiency in two half-siblings with a clinical diagnosis of TCS. After a diagnostic odyssey of almost a decade, multi-omics investigations finally resolved a SINE-VNTR-Alu (SVA) retrotransposon insertion in intron 17 of *TCOF1* leading to impaired production of the canonical *TCOF1* isoform. We confirmed the presence of the insertion in both affected individuals, and the inheritance from their mosaic healthy parent. We describe a type of variant not previously found in TCS and a novel pathogenic mechanism, emphasizing the utility of combined technologies to identify and interpret complex pathogenic variants currently missed by short-read exome sequencing.

## Results

We were referred a toddler (III-3) presenting with a clinical TCS phenotype, including mandibular and maxillary hypoplasia, microtia and hearing loss, down slanted palpebral fissures, cleft palate, and obstructive airway problems, requiring several surgical procedures (Fig. 1A & B). Routine diagnostics consisting of exome sequencing (ES) and chromosomal microarray analysis did not reveal pathogenic variants in *TCOF1, POLR1B, POLR1C*, or *POLR1D*, or in other Mendelian disease genes. The toddler remained genetically undiagnosed and passed away after a choking incident.

**Figure 1:**
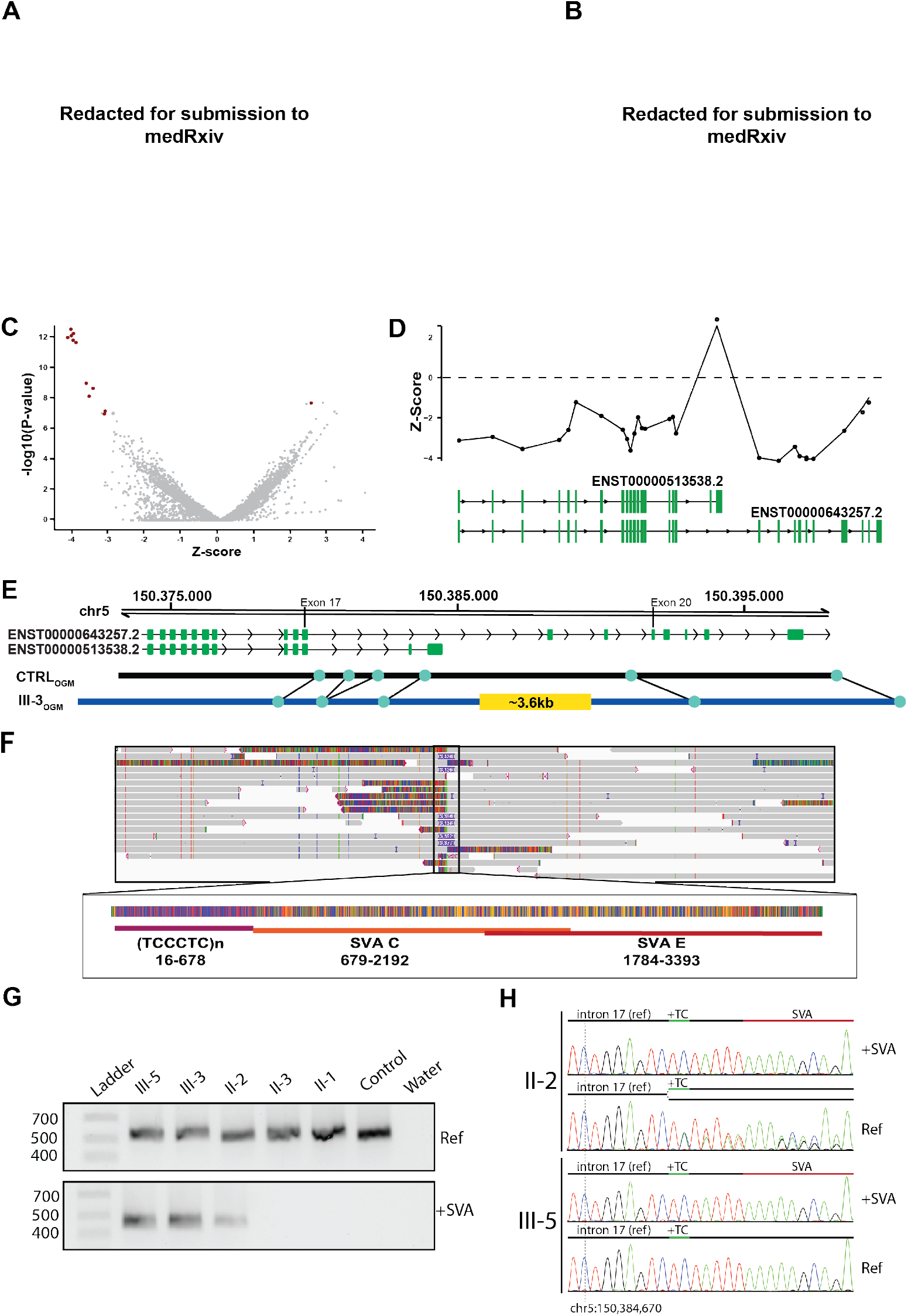
Identification of an intronic SVA insertion in *TCOF1* causing TCS. **A.** Pedigree including the affected individuals. Black arrow highlights the index of this study, asterisks indicate individuals whose DNA was available for the investigation. **B**. Photograph of III-3. Note severe mandibular hypoplasia/retrognathia, microtia and tracheostomy. **C**. Exon-level volcano plot showing the relative decreased expression of most of the *TCOF1* exons and upregulation of one of them (belonging to the isoform c) (red) vs. other genes’ exons (grey). **D**. Z-score plot showing the relative expression level of the exons of *TCOF1* long and short isoforms, drawn below. **E**. Schematic representation of the optical genome mapping results. **F**. RepeatMasker annotation results of the intronic SVA insertion. **G**. Gel electrophoresis of the allele-specific PCR products confirming the SVA insertion and showing its segregation in the pedigree. **H**. Sanger sequencing results of the allele-specific PCR products.

Years after the examination of III-3, an infant half-sibling (III-5), also with a clinical presentation strongly suggestive of TCS, was referred to us (Fig. 1A) but molecular genetic investigations as in III-3 could remained inconclusive. Subsequent short-read RNA-Seq (srRNA-Seq) outlier analysis in fibroblasts ^4^ from III-5 indicated reduced expression of the canonical *TCOF1* transcript (ENST00000643257.2) and apparent upregulation of a minor *TCOF1* transcript, isoform c ^5^(ENST00000513538.2) (Fig. 1C & D). Neither re-inspection of ES data nor targeted PCR and Sanger sequencing analysis identified variants that could explain the abnormal *TCOF1* expression. OGM of fibroblast DNA of III-5 suggested a heterozygous ~3,600 bp insertion between exon 17 and 20 of the canonical *TCOF1* transcript but could not resolve the insertion precisely (Fig. 1E).

Therefore, we performed nanopore long read genome sequencing (LR-GS) in DNA isolated from blood of III-5 and identified a heterozygous 3,396 bp (chr5(GRCh38):g.150384687_150384688ins3396) insertion in intron 17 of the canonical *TCOF1* isoform, just downstream of the minor *TCOF1* isoform c (Fig. 1F), unobserved in 48 unrelated in-house long-read genomes. The inserted sequence consisted of a 1) hexamer repeat (TCCCTC)n (nucleotides 16-678), 2) a retrotransposon of class SINE-VNTR-Alu (SVA) C (nucleotides 679-2192) and 3) a partially overlapping SVA E (nucleotides 1784-3393) (Fig. 1F). The hexamer repeat was determined to be part of the SVA C, as previously described ^6^. The 15 nucleotides upstream likely represent target site duplication, a characteristic hallmark of retrotransposon insertion ^7^.

We confirmed the insertion in DNA isolated from blood in both half-siblings and one parent with allele-specific PCR, agarose gel electrophoresis and Sanger sequencing (Fig. 1G).

Sanger sequencing of the PCR product without SVA from the unaffected parent revealed the presence of two alleles therein, marked by the heterozygous polymorphism chr5:150384675-G-GTC (gnomAD v4.1.0 minor allele frequency 60%) (Fig. 1H). This suggests that the unaffected parent is a gonadosomatic mosaic carrier of the SVA, explaining how they could transmit a pathogenic *TCOF1* variant while being clinically unaffected.

We then investigated how the SVA insertion could explain the isoform switch inferred from the srRNA-Seq data. SpliceAI^8^ predicted donor and acceptor sites within the SVA suggesting its possible exonization (Fig 2A). To investigate this, we included the insertion in the reference genome and realigned the srRNA-Seq data to the bespoke reference sequence. In both cycloheximide-treated and untreated cells, we detected the expression and incorporation of ~750 bp of the SVA-C together with *TCOF1* exons (Fig. 2B), in correspondence to the sites predicted by SpliceAI (Fig. 2A), confirming the SVA exonization, already described for other pathogenic SVA insertions^9^. Finally, to completely resolve the impact of the insertion on *TCOF1* transcripts, we performed RNA-Seq of native RNA from cycloheximide-treated fibroblasts of III-5 (Fig. 2A). We phased the reads and observed that while most of the reads from the wild-type allele represented the canonical isoform of *TCOF1* (ratio 29:1 long over short), the allele containing the SVA was preferentially represented by the isoform c (ratio 5:19 long over short)(Fig 2C). Moreover, the five reads representing the long isoform from the SVA-containing allele always contained. This was consistent with the short-read RNA-Seq data and highlighted a complex splicing pattern involving different exons and the SVA insertion. *In silico* translation of the reads containing the SVA revealed a frameshift and the introduction of a termination codon within the exonized SVA sequence likely to cause nonsense-mediated mRNAdecay^10^.

**Figure 2:**
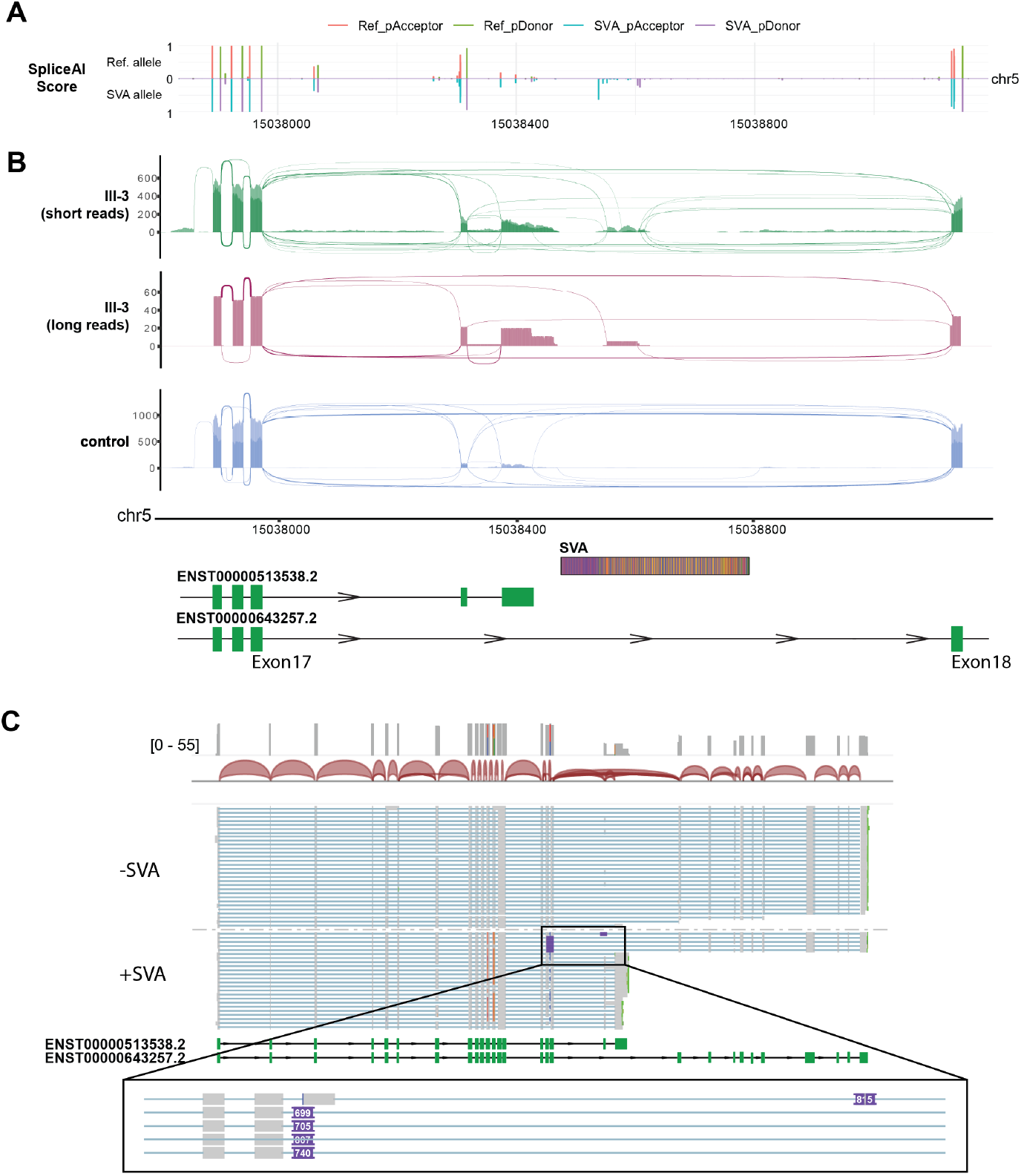
Functional characterization of the intronic SVA in TCOF1: **A.** SpliceAI predicted scores on reference allele and allele containing the SVA sequence. **B**. ggsashimi plot of the aligned short and long-read RNA-Seq data from III-5 and unrelated control on a reference genome containing the SVA sequence as detected in III-5. For short-read data, two overlapping tracks are provided; light colour is CHX-cells, darker color CHX+ cells. Schematics of the transcripts and of the SVA are aligned below the ggsashimi. **C**. IGV snapshot of the phased direct long-read RNA-Seq of fibroblasts from III-5.

## Discussion

We described two half-siblings with TCS, caused by the insertion of ~3.4 kb SVA-type retrotransposon into intron 17 of *TCOF1* transmitted by their mosaic unaffected parent. SVA insertions, as described here, have not been previously reported as a genetic cause for TCS nor to cause a pathogenic isoform switch. Noteworthy, pathogenic SVA insertions previously reported in other disorders belonged to the classes E and F ^11^ and this is the first instance of a hybrid E/C-class SVA associated with a genetic disease.

Multi-omics investigations were crucial in detecting *TCOF1* expression disruption, the pathogenic intronic SVA insertion, and in clarifying its impact. Given that predicted protein product of the short isoform lacks a nucleolar localization signal ^5^, it seems likely that a switch to this isoform would abolish the role of TCOF1 in ribosome biosynthesis, converging on the same molecular mechanism as other TCS-causing variants, so far only reported for the long canonical isoform.

The accurate mapping and identification of transposable element insertions remain a significant challenge both for SR-GS ^12^ and for srRNA-Seq. Specific tools aimed at addressing these challenges are emerging and, while they are not yet established for diagnostic purposes, they will likely enhance the diagnostic yield of similar events while also determining their consequences at the mRNA level.

Retrotransposons have been increasingly recognized as causative events in genetic diseases ^13,14^. Given that *de novo* transposition events have been estimated to occur at rates of up to 1:20 live births for just L1 elements alone^15^, retrotransposon-mediated gene disruption could be more common than currently recognized due to technological limitations.

Strikingly, both half-siblings inherited the SVA insertion from their healthy parent, who carries the insertion in mosaic form, suggesting that the SVA activation and retrotranscription occurred during early embryonic development and is not limited to only gametogenesis. Interestingly, a family with a similar inheritance pattern but with a pathogenic frameshift variant was reported recently ^16^. Given that TCS is considered to exhibit incomplete penetrance and 60 % of cases are thought to result from *de novo* mutations, we suspect that there are more mosaic parental carriers that account for part of this.

In summary, by combining several innovative molecular diagnostic techniques, we identified an intronic SVA insertion disrupting *TCOF1* in a family with TCS. In addition to providing the family with a molecular diagnosis, in this case understanding the inheritance was also directly relevant for the reproductive choices of the siblings of the carrier parent. Our results demonstrate the added value of combining different technical approaches for variant detection.

## Materials & Methods

Informed consent for diagnostic tests, research investigations, to share photograph, clinical, and analysis data was obtained. Chromosomal microarray and ES were performed following standard operating procedures^17^. Use of genome-wide technologies for diagnostic purposes was previously approved (Institutional review board MEC-2012-387).

srRNA-Seq was performed as recently described ^4^. For nanopore direct long-read RNA-Seq, RNA was performed following the Oxford Nanopore Nanopore Direct RNA sequencing (SQK-RNA004) protocol.

Genomic DNA was extracted following standard operating procedures. OGM was conducted as previously described^18^. For LR-GS, gDNA was processed following the Oxford Nanopore Ligation Sequencing Kit (SQK-LSK114). Basecalling and alignment to the reference genome were performed in MinKNOW v23.07.12. Structural variants were called by sniffles2 ^19^. The SVA insertion was annotated with RepeatMasker v4.1.5 ^20^.

A universal forward primer (5’-TCTCCACAACAGCCCTATGA-3’) was used in combination with a reverse primer specific for the allele with (5’-GAGACGGGACCTTTTCTGC-3’) or without the insertion (5’-AAAAATCTCTCAATCAGGAAGAGG-3’) with expected product size of 420 bp and 498 bp, respectively.

## Data Availability

All data produced in the present work are contained in the manuscript

## Acknowledgements

We thank the patients and families for their participation.

## Author contributions

Conceptualization: F.F., M.D., M.v.D., T.v.H.;

Methodology: F.F., M.N., D.R., L.v.U., B.M., H.C.v.d.L., R.S., M.K.;

Formal analysis and investigation: All authors

Writing - original draft preparation: N.K., F.F., M.D., T.v.H.;

Writing - review and editing: N.K., M.D., F.F., D.R., H.C.v.d.L., T.S.B., M.N., T.K., M.W., M.v.D., T.v.H.;

Supervision: T.K., T.v.H.

All authors read, commented, and approved the final manuscript.

